# Unravelling undiagnosed rare disease cases by HiFi long-read genome sequencing

**DOI:** 10.1101/2024.05.03.24305331

**Authors:** Wouter Steyaert, Lydia Sagath, German Demidov, Vicente A. Yépez, Anna Esteve-Codina, Julien Gagneur, Kornelia Ellwanger, Ronny Derks, Marjan Weiss, Amber den Ouden, Simone van den Heuvel, Hilde Swinkels, Nick Zomer, Marloes Steehouwer, Luke O’Gorman, Galuh Astuti, Kornelia Neveling, Rebecca Schüle, Jishu Xu, Matthis Synofzik, Danique Beijer, Holger Hengel, Ludger Schöls, Kristl G Claeys, Jonathan Baets, Liedewei Van de Vondel, Alessandra Ferlini, Rita Selvatici, Heba Morsy, Marwa Saeed Abd Elmaksoud, Volker Straub, Juliane Müller, Veronica Pini, Luke Perry, Anna Sarkozy, Irina Zaharieva, Francesco Muntoni, Enrico Bugiardini, Kiran Polavarapu, Rita Horvath, Evan Reid, Hanns Lochmüller, Marco Spinazzi, Marco Savarese, Solve-RD DITF-ITHACA, Solve-RD DITF-Euro-NMD, Solve-RD DITF-RND, Solve-RD DITF-EpiCARE, Leslie Matalonga, Steven Laurie, Han G. Brunner, Holm Graessner, Sergi Beltran, Stephan Ossowski, Lisenka E.L.M. Vissers, Christian Gilissen, Alexander Hoischen, the Solve-RD consortium

## Abstract

Solve-RD is a pan-European rare disease (RD) research program that aims to identify disease-causing genetic variants in previously undiagnosed RD families. We utilised 10-fold coverage HiFi long-read sequencing (LRS) for detecting causative structural variants (SVs), single nucleotide variants (SNVs), insertion-deletions (InDels), and short tandem repeat (STR) expansions in extensively studied RD families without clear molecular diagnoses. Our cohort includes 293 individuals from 114 genetically undiagnosed RD families selected by European Rare Disease Network (ERN) experts. Of these, 21 families were affected by so-called ‘unsolvable’ syndromes for which genetic causes remain unknown, and 93 families with at least one individual affected by a rare neurological, neuromuscular, or epilepsy disorder without genetic diagnosis despite extensive prior testing.

Clinical interpretation and orthogonal validation of variants in known disease genes yielded thirteen novel genetic diagnoses due to *de novo* and rare inherited SNVs, InDels, SVs, and STR expansions. In an additional four families, we identified a candidate disease-causing SV affecting several genes including an *MCF2*/*FGF13* fusion and *PSMA3* deletion. However, no common genetic cause was identified in any of the ‘unsolvable’ syndromes. Taken together, we found (likely) disease-causing genetic variants in 13.0% of previously unsolved families and additional candidate disease-causing SVs in another 4.3% of these families.

In conclusion, our results demonstrate the added value of HiFi long-read genome sequencing in undiagnosed rare diseases.

## Introduction

Rare diseases (RD) affect 400 million people worldwide (Nguengang Wakap et al., 2020). It is estimated that 80% of these diseases have a genetic origin (Sernadela et al., 2017). Pinpointing the disease-causing genetic variant is important for RD families because it ends an often time-consuming, stressful, and costly diagnostic odyssey (Biesecker & Green, 2014). In addition, several disease management strategies and treatment options depend on the specific disease gene or variant (Pogue et al., 2018).

With routinely used short-read sequencing (SRS) technologies, such as exome and genome sequencing, diagnostic yields vary between 8% and 70%, depending on the diseases studied and inclusion criteria used (Wright et al., 2018). Recently, a large-scale reanalysis effort of exomes and genomes from undiagnosed disease families has been conducted within the Solve-RD consortium; this study illustrates that updated knowledge and improved variant identification and interpretation substantially increase diagnostic yield (S. Laurie, W. Steyaert, E. de Boer et al., Nat Med in revision). Other re-analysis efforts have resulted in similar increases in diagnostic yield (Bullich et al., 2022; Liu et al., 2019; Wright et al., 2018). Despite this, most tested RD patients remain without a genetic diagnosis. Besides incomplete knowledge of the functional and phenotypic consequence of genetic variation, shortcomings at the variant identification level substantially contribute to the fact that many RD patients remain genetically undiagnosed. Indeed, SRS technologies result in an almost complete characterisation of short genetic variants (single and multi-nucleotide substitutions and small insertions and deletions) in the unique portions of an individual’s genome, but the analysis of duplicated and repetitive genomic regions and particularly the identification of structural variants (SVs) and short tandem repeat (STR) expansions remain far from complete (Chaisson et al., 2019; Chintalaphani et al., 2021; Porubsky et al., 2023).

Several recent studies demonstrate that long-read sequencing (LRS) technologies uncover a whole new reservoir of (structural) genetic variation (Beyter et al., 2021; Chaisson et al., 2019; Harvey et al., 2023; Kucuk et al., 2023; Pauper et al., 2021; Zook et al., 2020). This is especially true for SVs of intermediate size (50 to a couple of thousand base pairs), which are not identified in SRS data, nor with molecular cytogenetic technologies such as multiplex-ligation dependent probe amplification (MLPA) or comparative genomic hybridisation arrays (aCGH). Now that these LRS technologies produce high-quality sequencing reads at steadily dropping costs, researchers can evaluate the hypothesis that part of the genetically undiagnosed RDs are caused by variants that remain hidden from previously used technologies. The exploration and interpretation of SVs in undiagnosed RD families has indeed shown to be successful in the past couple of years for several disease phenotypes (Fadaie et al., 2021; Merker et al., 2018; Mizuguchi, Suzuki, et al., 2019; Mizuguchi, Toyota, et al., 2019; Sabatella et al., 2021; Sanchis-Juan et al., 2018; Zeng et al., 2019).

Here, as part of the Solve-RD consortium effort, we applied long-read genome sequencing to two unique patient cohorts. Firstly, a cohort of 21 families (including 16 trios) with clinically well-recognized, so-called ‘unsolvable’ syndromes. These syndromes have been extensively studied by many groups using various genomic technologies over many years, but the genetic causes for them remain unknown. The second cohort consisted of 232 individuals from 93 families with rare neurological, neuromuscular, or epilepsy disorders. While most of these patients are affected by conditions for which several genetic causes are known, these particular families remained ‘unsolved’; as extensive diagnostic and/or research testing, including prior exome or genome sequencing, had failed to yield a diagnosis.

## Results

We analysed the genomes of 293 individuals from 114 previously undiagnosed RD families using HiFi long-read sequencing (Fig. 1; Supplemental Table S1). Part of the cohort consists of families selected by experts from ERN-ITHACA - mostly parent-offspring trios - with a clinically well-recognizable syndrome termed ‘unsolvable’ syndromes (n=21), including Aicardi (MIM ID %304050), Hallermann-Streiff (%234100), Gomez-Lopez-Hernandez (%601853), Pai (%155145), and syndromes belonging to the oculoauriculovertebral spectrum, all of which remain genetically elusive despite huge global efforts to identify the disease cause. The other part of the cohort (n = 93) are RD families with a rare neurological, neuromuscular, or epilepsy disorder, selected by experts from ERN-EURO-NMD, ERN-EpiCARE, ERN-RND, and ERN-ITHACA.

**Figure 1:**
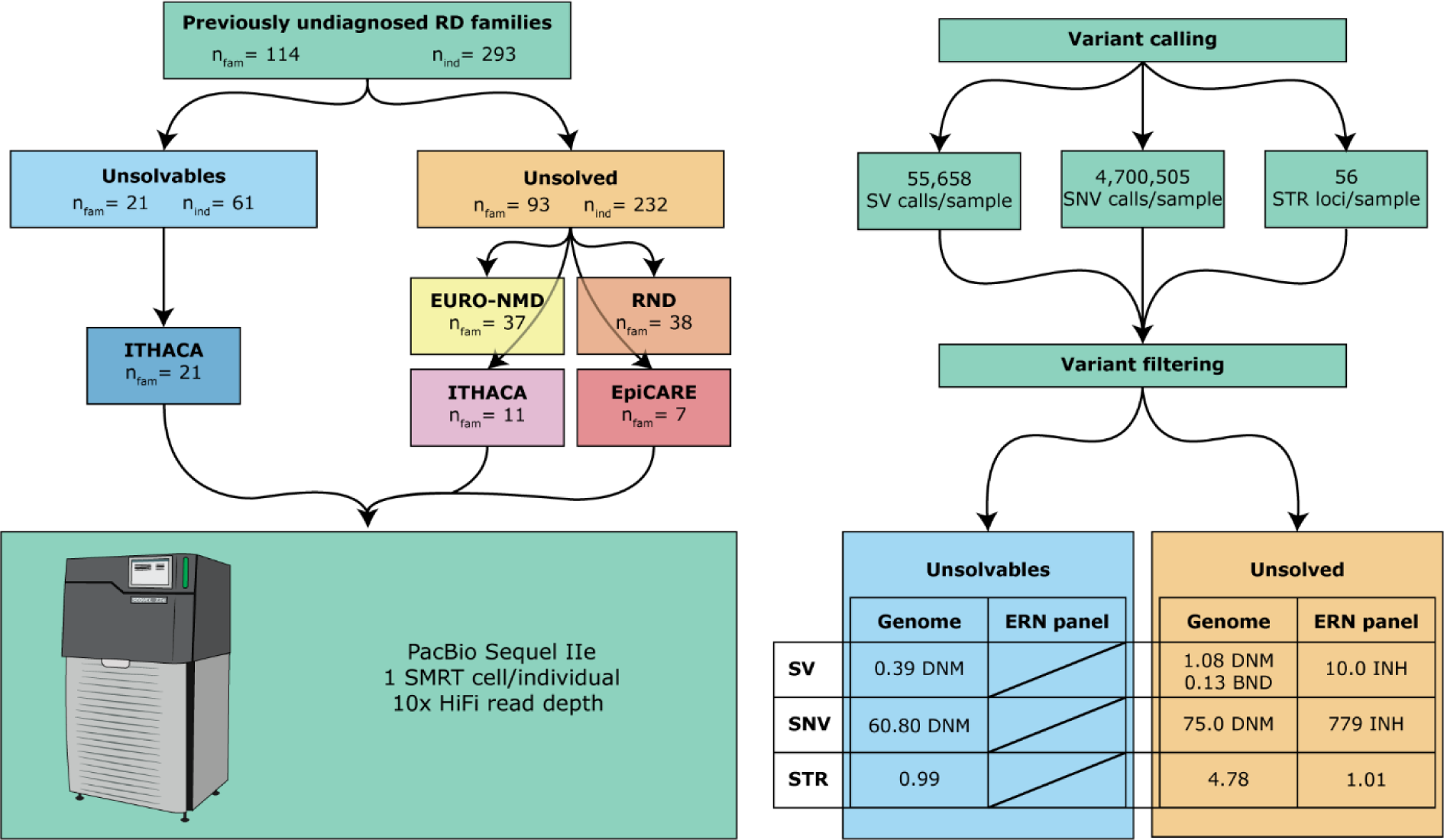
HiFi long-read sequencing in a unique cohort of 293 individuals from 114 RD families. The study cohort consists of two subcohorts: the ‘unsolvables’ (families affected by clinically well-recognizable syndromes for which the cause is yet unknown) and the ‘unsolved’ (families affected by a rare neurological, neuromuscular or epilepsy disease). All patients were recruited via four European Reference Networks and subsequently sequenced using a single SMRT cell of sequencing data per individual. Genome-wide calling of SVs and SNVs was conducted and STRs were genotyped at 56 known disease-associated loci.

All of these families and syndromes were genetically well-studied in the past using SRS and other applicable approaches but without diagnostic success. Consequently, we hypothesized that part of these syndromes are caused by genetic variants, mostly SVs or STRs, that cannot be identified using SRS or probe-based technologies. Due to the lack of effective population databases for SVs, we focused our study on *de novo* events in parent-offspring trios, on large inherited SVs (> 100 kb), and SVs affecting a disease-relevant gene, since these events are more likely to affect the phenotype (Coe et al., 2014). We also genotyped 56 known disease-associated STR loci since these loci are highly relevant for neurological disease, yet difficult to characterize using SRS techniques. To fully exploit our sequencing data, we also identified and filtered SNVs and InDels.

On average, we identified 55,658 SVs (≥ 20 bp; 23,385 SVs ≥ 50 bp) and 4,700,505 SNVs per individual. Of these, 13,481 SVs and 43,172 SNVs are private to one family (Methods; Supplemental Table S2). From the 18 visually curated putative *de novo* SVs for which flanking sequencing primers could be designed, four were confirmed as *de novo* variants in the child. In turn, five calls were false positives, six of the variants turned out to be true but inherited from a parent and 3 other variants were true positives too but the parental sequences failed (Methods; Supplemental Fig. S1; Supplemental Table S3).

### Identification of (likely) pathogenic variants in previously undiagnosed RD

#### Unsolvable syndromes

In the subcohort consisting of 21 families with ‘unsolvable’ syndromes, we could not identify a gene or locus in which rare (*de novo*) variants were present in multiple families with the same syndrome. However, in a sporadic female patient (F036.1) initially diagnosed with Aicardi syndrome, we identified a *de novo* missense variant in *TUBA1A* (Tubulin alpha 1a, MIM ID *602529, NM_006009.4, chr12:g.49,185,725C>T, c.641G>A, p.(Arg214His); Figure 2; Table 1). The variant has previously been described as a cause of lissencephaly 3 (LIS3, MIM ID #611603) (Bahi-Buisson et al., 2014a). Clinical reassessment of the patient’s phenotype confirmed the new diagnosis.

**Figure 2:**
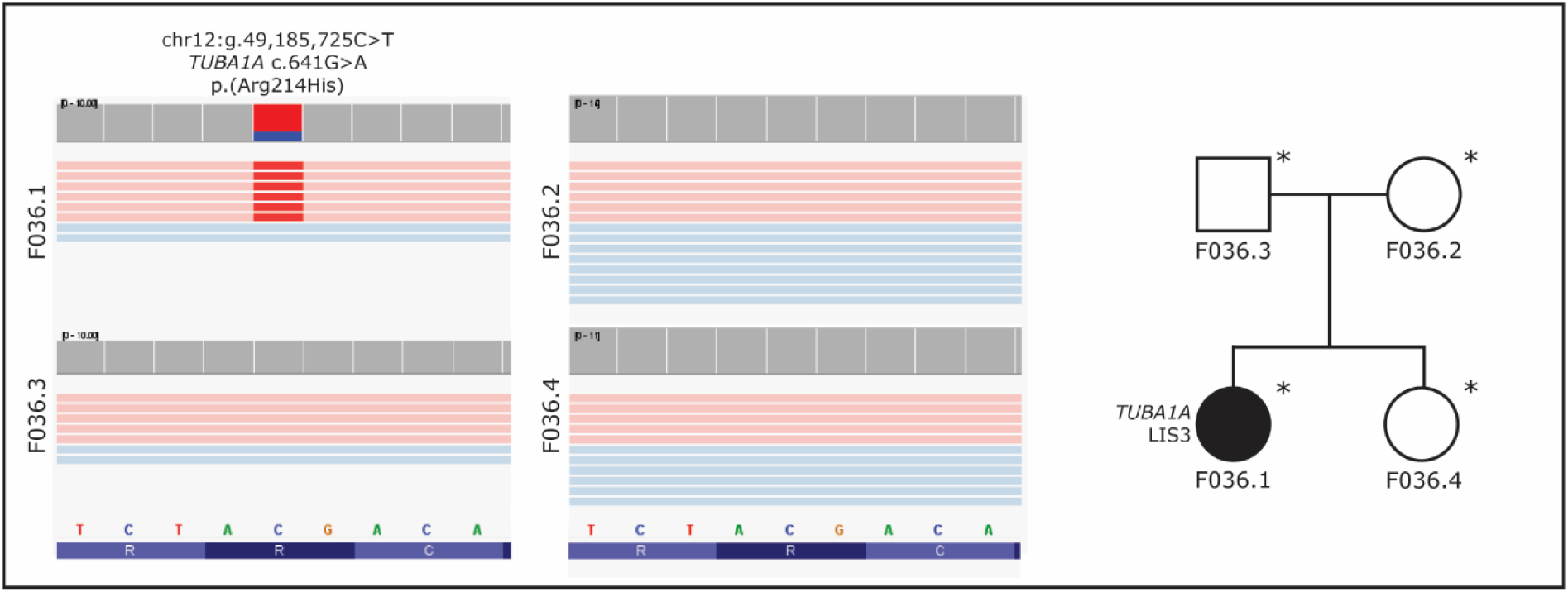
Visualization of the *TUBA1A de novo* missense variant in F036.1 using IGV, and a pedigree of the family. The variant has earlier been described as a cause of lissencephaly. The healthy family members do not carry the variant. Sequenced individuals are marked with an asterisk (*) in the pedigrees.

**Table 1:** Overview of disease-causing (DC) and candidate disease-causing (cDC) genetic variants in the complete study cohort.

| Participant ID | ERN | Cohort | DC/cDC | Gene name(s) | Variant type | Orthogonal validation |
| --- | --- | --- | --- | --- | --- | --- |
| F036.1 | ITHACA | Unsolvable | DC | *TUBA1A* | (*de novo*) SNV | - |
| F034.1 | EURO-NMD | Unsolved | DC | *DMD, NHS* | SV (inversion) | Sanger |
| F043.1 | EURO-NMD | Unsolved | DC | *DMD* | SV (inversion) | OGM |
| F008.1 | RND | Unsolved | DC | *NOP56* | STR | RNA-Seq |
| F049.1 | RND | Unsolved | DC | *NOP56* | STR | RNA-Seq |
| F047.1 | RND | Unsolved | DC | *DAB1* | STR | - |
| F051.1 | RND | Unsolved | DC | *RFC1* | STR | RNA-Seq |
| F071.1 | EURO-NMD | Unsolved | DC | *DMD* | (deep intronic) SNV | Sanger |
| F084.1 | RND | Unsolved | DC | *SPAST* | (intronic) SNV | ES, exon-skipping, Sanger |
| F044.1 | RND | Unsolved | DC | *TTN* | (*de novo*) SNV | SRS |
| F112.1 | EURO-NMD | Unsolved | DC | *MYOT* | SV (tandem duplication) | SRS |
| F087.1 | EURO-NMD | Unsolved | DC | *REEP1* | (deep intronic) SNV | - |
| F016.1 | RND | Unsolved | DC | *REEP1* | SV (deletion) | PCR + LRS |
| F039.1 | EURO-NMD | Unsolved | cDC | *FGF13, MCF2 and F9* | (*de novo*) SV (duplication) | PCR + LRS + cDNA + RNA-seq |
| F075.1 | EURO-NMD | Unsolved | cDC | *PSMA3* | (*de novo*) SV (deletion) | PCR + Sanger |
| F065.1 | ITHACA | Unsolved | cDC | *CPE, TLL1, NEK1, CLCN3, ...* | SV (5Mb duplication) | Array CGH, ES |
| F022.1 | RND | Unsolved | cDC | *ARMC9* | SV (300 kb duplication) | - |

### Disease-causing variants identified in rare neurological, neuromuscular and epilepsy diseases

After prioritizing and clinically interpreting genetic variants in the 93 families from the ‘unsolved’ cohort, we established a genetic diagnosis in 12 of them (Methods; Table 1).

#### Large SVs

In two unrelated male patients (F043.1 and F034.1; Fig. 3A-D) with muscular dystrophy, we identified disease-explanatory inversions breaking *DMD* (Dystrophin, MIM ID *300377; Fig. 3A, 3C). The breakpoints in patient F43.1 are chrX:23,308,848 and chrX:32,004,110 (hg38) resulting in an inversion of 8.7 Mb which breaks *DMD* in intron 44 (NM_004006.3), resulting in a truncated transcript (Fig. 3A). This event was initially discovered by optical genome mapping (OGM) and LRS detected the exact breakpoints of the event. In patient F034.1 an inversion of chrX:17,398,320-32,130,845 was identified (Fig. 3C). This event breaks *NHS* in intron 1 (NM_0129186.2) and *DMD* in intron 44 (NM_004006.3). This event was confirmed by Sanger sequencing, which also highlighted the insertion of a short ATAAT sequence in the first intron of *NHS*, and of a 38-nucleotide sequence in the intron 44 of *DMD*, which likely favoured the inversion. As these genes have opposite orientations on the chromosome, the inversion results in two theoretical fusion genes in which the exon orientation is conserved. However, neither theoretical gene product is in frame much past the fusion breakpoint. The likely disruption of both genes is also in line with the patient’s phenotype, who in hindsight presents not only with a dystrophy but also with a cataract which is a characteristic feature of Nance-Horan syndrome (MIM ID #302350) caused by loss-of-function variants of *NHS*.

**Figure 3:**
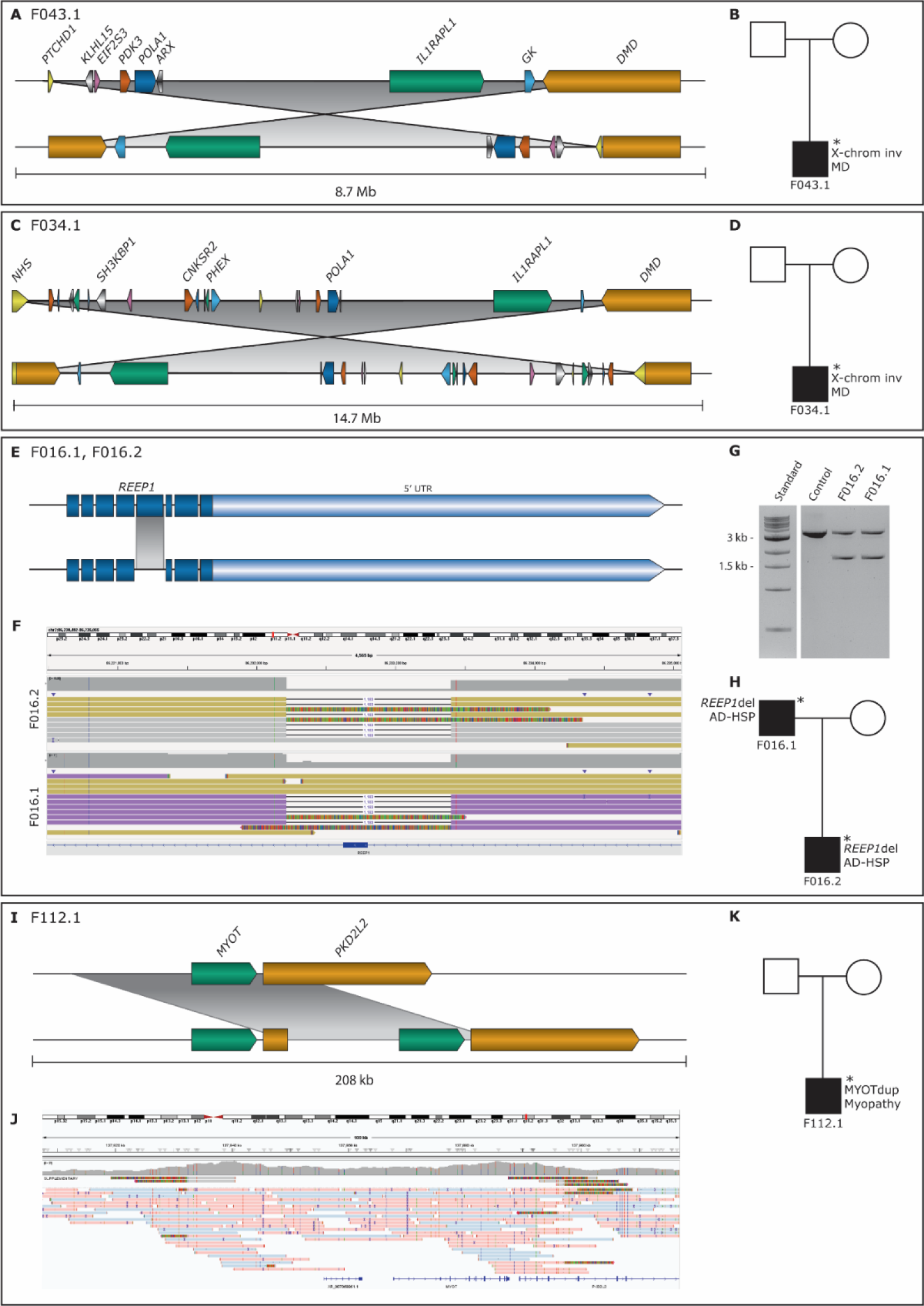
Visualization of disease-causing SVs in the ‘unsolved’ cohort in the form of cartoons and/or IGV screenshots, along with corresponding pedigrees. In two unrelated male patients (F043.1 in A and B, F034.1 in C and D) with muscular dystrophy, we found X-chromosomal inversions (A-D). In both cases, *DMD* is disrupted (A and C), in one a second gene disruption adds to the phenotype (C). In a father and son with hereditary spastic paraplegia, we detected a deletion of *REEP1* exon 6 (E-H). The deletion in F016.1 and F016.2 is shown here as a cartoon (E) and as a screenshot in IGV (F). The deletion was also visualized by agarose gel electrophoresis, which confirms that both patients are heterozygous for the deletion (G). The pedigree of the family is shown in (H). In a patient with adult-onset distal myopathy, a 65 kb duplication involving *MYOT* (I) was confirmed to be in tandem by LRS (J). The pedigree of the family is shown in K. Sequenced individuals are marked with an asterisk (*) in the pedigrees (B, D, G, H, K). Abbreviations: MD = muscular dystrophy, AD-HSP = autosomal dominant hereditary spastic paraplegia.

In a duo consisting of an affected father and affected son (F016.1 and F016.2, respectively; Fig. 3E-H) presenting with hereditary spastic paraplegia, we detected a 1.2 kb deletion encompassing the entire exon 6 of *REEP1* (Receptor expression-enhancing protein 1, MIM ID *609139, NM_001371279.1). Both the father and the son are heterozygous carriers of this deletion (Fig. 3F-G). *REEP1* is expressed in larger degrees in all brain tissues, the gastrointestinal tract, and arterial tissues, but has some expression in most other tissue types as well. It regulates lipid droplet formation and the morphology of the endoplasmic reticulum (Renvoisé et al., 2016). Variants in *REEP1* have been described in autosomal recessive distal hereditary neuronopathy (MIM ID #620011) and autosomal dominant spastic paraplegia 31 (MIM ID #610250). Among known variants causing SPG31 are single-exon deletions in *REEP1* of exons 2 and 3 (Goizet et al., 2011). Additionally, two cases with deletions encompassing more than one exon have been described (Battini et al., 2011; Ishiura et al., 2014), neither affecting exon 6.

In a singleton patient with adult-onset distal myopathy (F112.1, Figure 3I-K), a 65kb duplication involving *MYOT* (myotilin, MIM ID *604103, chr5:137,832,296-137,897,203) had earlier been identified by a gene panel for myofibrillar myopathy. The orientation of the duplication was uncertain but was confirmed to be in tandem by LRS. Heterozygous variants in *MYOT* are a known cause of myofibrillar myopathy 3 (MIM ID #609200), a slowly progressive muscle disorder with adult onset.

#### Repeat expansions

In two families with autosomal dominant (AD) ataxia, we identified disease-explanatory heterozygous expansions of the GGCCTG motif in intron 1 of the *NOP56* gene (Table 1; NOP56 ribonuclear protein, NM_006392.4, MIM ID *614154). Repeat expansions in *NOP56* are a known cause of AD spinocerebellar ataxia 36 (SCA36, MIM ID #614153) (Kobayashi et al., 2011). The hexanucleotide motif count in a duo consisting of two affected siblings (F008.1 and F008.2; Fig. 4A) was estimated at >1200. In the other family, consisting of two affected family members in two generations and one unaffected family member (F049.1, F049.2, and F049.3, respectively, Fig. 4B), the motif count was >34 in the affected mother and >45 in the affected child. The pathogenic repeat threshold of NOP56 is generally regarded to be 650 hexanucleotide repeats, however, shorter repeats are also known to be causative (Obayashi et al., 2015). The repeat expansion in the latter family was also discovered by reduced expression through RNA-sequencing and whole genome sequencing by parallel efforts in Solve-RD.

**Figure 4:**
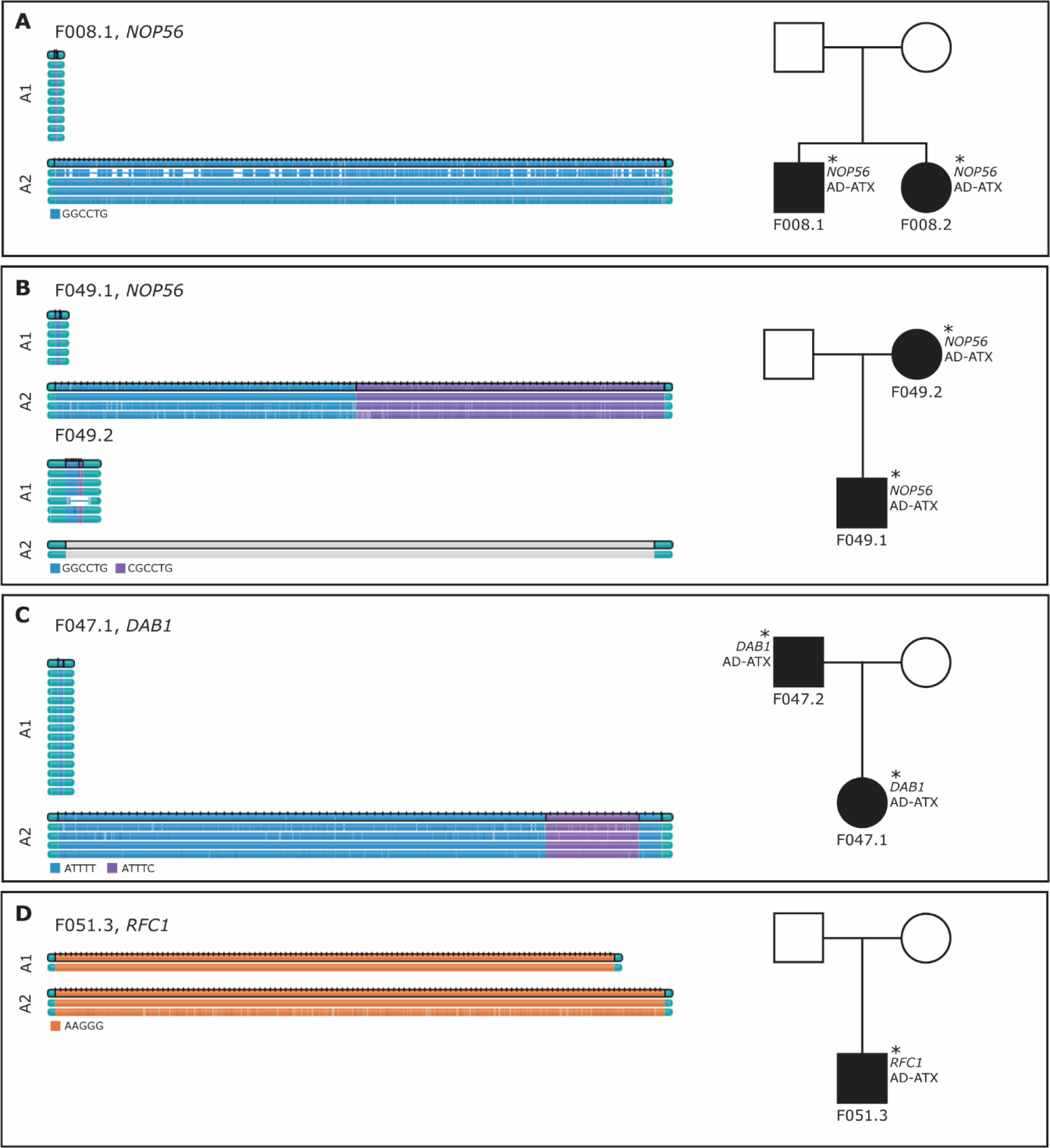
Visualizations produced using the PacBio TRGT tool and pedigrees for the families with pathogenic STR expansions. In siblings F008.1 and F008.2, a heterozygous GGCCTG expansion in *NOP56* was detected (A). In another family, an expansion including the motifs GGCCYG and CGCCTG in *NOP56* was detected in one generation (F049.1), and the STR expansion was subsequently also identified in the mother (B). In patient F047.1 and their father, we identified heterozygous STR expansion *DAB1* including both ATTTT and ATTTC motifs (C). In another patient, we identified homozygous STR expansions in *RFC1*. Alleles are denoted by “A1” and A2”. Sequenced individuals are marked with an asterisk (*) in the pedigrees. Abbreviations: AD-ATX = autosomal dominant ataxia.

In a family presenting with autosomal dominant ataxia, we found a repeat expansion in *DAB1* (MIM ID *603448), a known causal gene for Spinocerebellar ataxia 37 (SCA37, MIM ID #615945). Age-dependent penetrant alleles have been reported to have an insertion of 31-75 ATTTC repeats, while the normal motifs are usually uninterrupted and consist of 7-400 units of ATTTT (Matilla-Dueñas & Volpini 2019). The analysis of LRS data indicated the presence of two alleles in the index case F047.1, one with 7 ATTTT repeats and another with a complex structure of (estimated) 615 ATTTT motifs, followed by 117 ATTTC repeats and then again by 29 ATTTT repeats (Fig. 4C), supported by 5 high quality spanning PacBio reads.

Finally, a homozygous repeat of the pathogenic AAGGG motif in the *RFC1* gene (MIM ID *102579) was found in a patient with ataxia (F051.3). Repeat expansions in *RFC1* are known to cause CANVAS-spectrum disorder (“Cerebellar ataxia, neuropathy, and vestibular areflexia syndrome”, MIM ID #614575), being very consistent with the observed phenotype. The number of AAGGG motifs was estimated by the tool to be 271 on one allele and 1181 on the other allele (Fig. 4D). However, it is possible that the first allele is longer than 271 pathogenic repeats, since it was inferred based on soft clipped reads, not reads spanning the full repeat. The visualization of LRS data from this patient in Integrative Genome Viewer (IGV) indicated that no normal alleles were present. The further validation of this likely causative repeat is prepared to be described elsewhere.

#### SNVs

In a sporadic male patient with suspected titinopathy (F071.1), presenting with progressive proximal muscle weakness, and myopathic features in his muscle biopsy, we identified a deep intronic SNV in *DMD* (chrX:33,174,335C>T) (Fig. 5A). This variant has previously been shown to be a cause of Becker muscular dystrophy (BMD, MIM ID #300376) through exonisation of a 149 bp sequence within intron 1 of *DMD* (Okubo et al., 2020). Clinical reassessment of the patient’s phenotype confirmed the BMD diagnosis.

**Figure 5:**
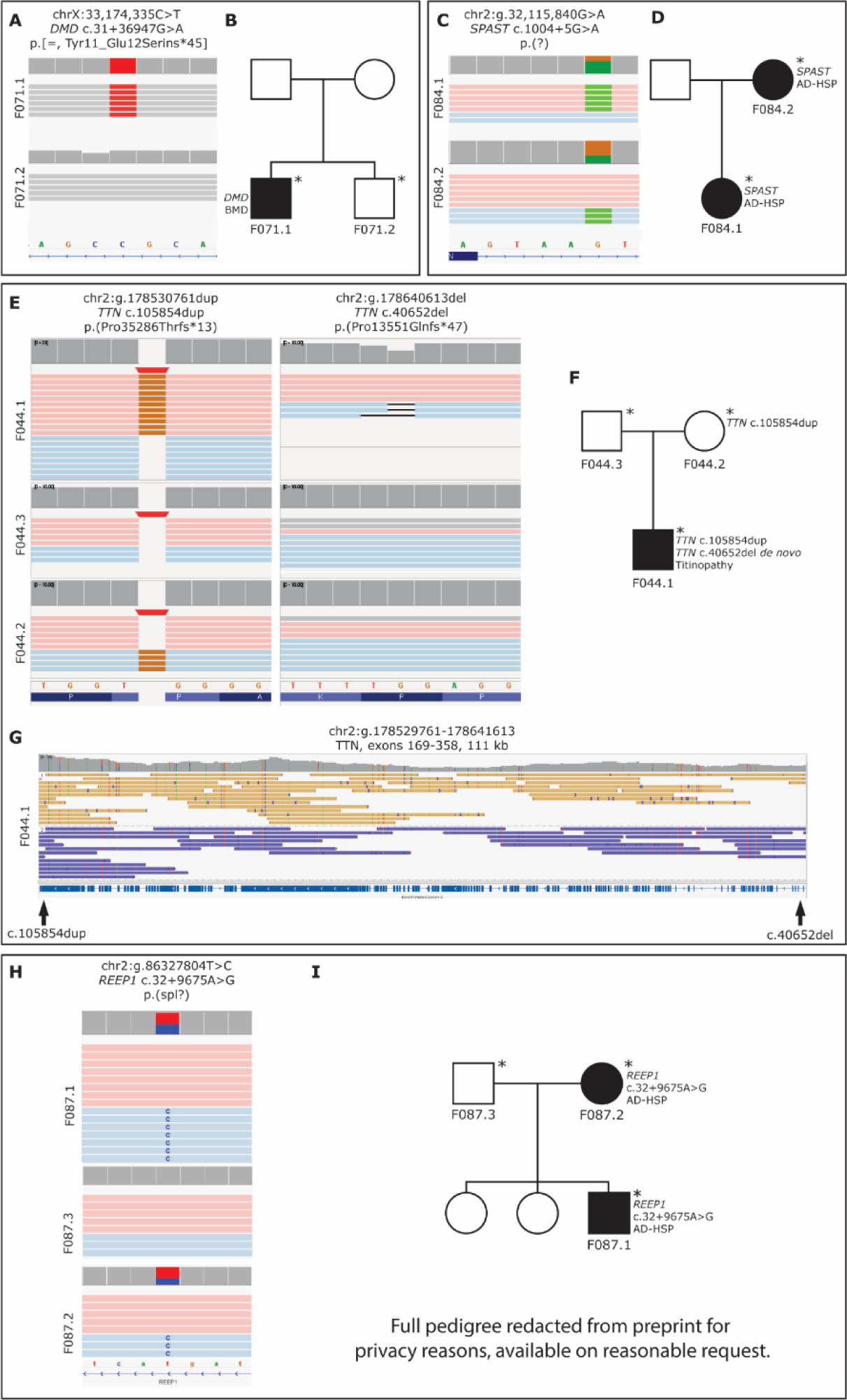
Visualization of disease-causing SNVs and InDels in the ‘unsolved’ cohort in the form of IGV screenshots, along with corresponding pedigrees. In a sporadic patient with suspected titinopathy (F071.1), we identified a deep-intronic variant in DMD (A). The nonaffected sibling (F071.2) did not carry the variant (A and B). In a duo consisting of an affected daughter and mother (F084.1, F084.2) with hereditary spastic paraplegia, we identified a non-canonical splice site variant in *SPAST* (C and D). In a patient with titinopathy (F044.1), a maternally inherited and a *de novo* variant had been identified earlier (C-F). The two variants are located 109 kb apart, but the alleles were successfully phased through the entire region by LRS (G). In a family with an affected mother and son (F084.1, F084.2), we detected an intronic variant in *REEP1* (H). The index patient also has an affected uncle, whose sample was not sequenced (I). The reads are coloured by haplotag; pink and light blue, or yellow and purple represent different alleles in A, C, E, G, and H. Unphased reads, such as X-chromosomal reads in males, are shown in grey (A and E). Sequenced individuals are marked with an asterisk (*) in the pedigrees (B, D, F, I). Abbreviations: BMD = Becker Muscular Dystrophy, AD-HSP = autosomal dominant hereditary spastic paraplegia.

In a duo consisting of an affected daughter and mother (F084.1, F084.2) with AD hereditary spastic paraplegia (HSP), we identified a variant in intron 6 of *SPAST* (Spastin, MIM ID *604277, NM_014946.4, chr2:g.32,115,840G>A, c.1004+5G>A) (Fig. 5B). Variants in *SPAST* are known to cause HSP4 (MIM ID #182601) (Hazan et al., 1999) and while the same variant has not been previously recorded, a variant affecting the same base has previously been evaluated as pathogenic in ClinVar (variation ID 989101). The variant was identified in parallel by the referring laboratory but was initially considered to be of uncertain significance. Subsequent RNA analysis eventually demonstrated skipping of exon 6 showing that the variant is likely pathogenic through loss-of-function exon-skipping.

In a sporadic patient with suspected titinopathy (F044.1), two pathogenic frameshift variants in *TTN* (titin, MIM ID *188840, NM_001267550.2) had been previously identified before submission to the Solve-RD collection. Of these, one was maternally inherited, chr2:g.178530761dup, c.105854dup, p.(Pro35286Thrfs*13), and one *de novo*, chr2:g.178640613del, c.40652del, p.(Pro13551Glnfs*47) (Fig. 5C). Both variants are located in ubiquitously expressed exons; the maternally inherited variant affects the constitutional exon 308, and the *de novo* variant affects exon 221, which is expressed in 99% of *TTN* transcripts (Savarese et al., 2018). Previous SRS efforts had not been successful in identifying on which allele the *de novo* event had occurred. Using our approach, we were able to successfully differentiate between the alleles and confirmed the two frameshift variants to be in trans, thus explanatory for the patient’s phenotype (Fig. 5D).

In a family with suspected autosomal dominant hereditary spastic paraplegia (AD HSP), we identified a deep intronic substitution in the first intron of *REEP1* (chr2:g.86327804T>C, NM_001371279.1:c.32+9675A>G), segregating in the affected mother and son (F087.1, F087.2). Previous genetic analysis with HSP and hereditary neuropathy panels was negative. The variant is predicted to alter splicing by activation of a cryptic donor site by Human Splicing Finder and MaxEntScan (Desmet et al., 2009; Yeo & Burge, 2003). Loss-of-function variants including splice-altering intronic variants in *REEP1* have previously been reported as causative in AD-HSP families (Züchner et al., 2006).

### Candidate disease-causing variants identified in rare neurological, neuromuscular and epilepsy diseases

In addition to the pathogenic variants identified above, in which the disease gene is well established and fits the patient’s phenotype according to clinical experts, our analyses revealed novel, likely pathogenic aberrations in four additional families (Table 1).

#### *De novo* X-chromosomal duplication

In a female patient (F039.1) presenting with arthrogryposis multiplex congenita, thoracolumbar scoliosis, and restrictive ventilatory defect, we discovered a 500 kb X-chromosomal tandem duplication, which was confirmed *de novo* in the patient by gel electrophoresis and sequencing (Fig. 6A-D). The breakpoints of the duplication disrupt two genes; *FGF13* (fibroblast growth factor 13, MIM ID *300070, NM_004114.5), and *MCF2* (Cell line-derived transforming sequence, *311030, NM_001171876.2).

**Figure 6:**
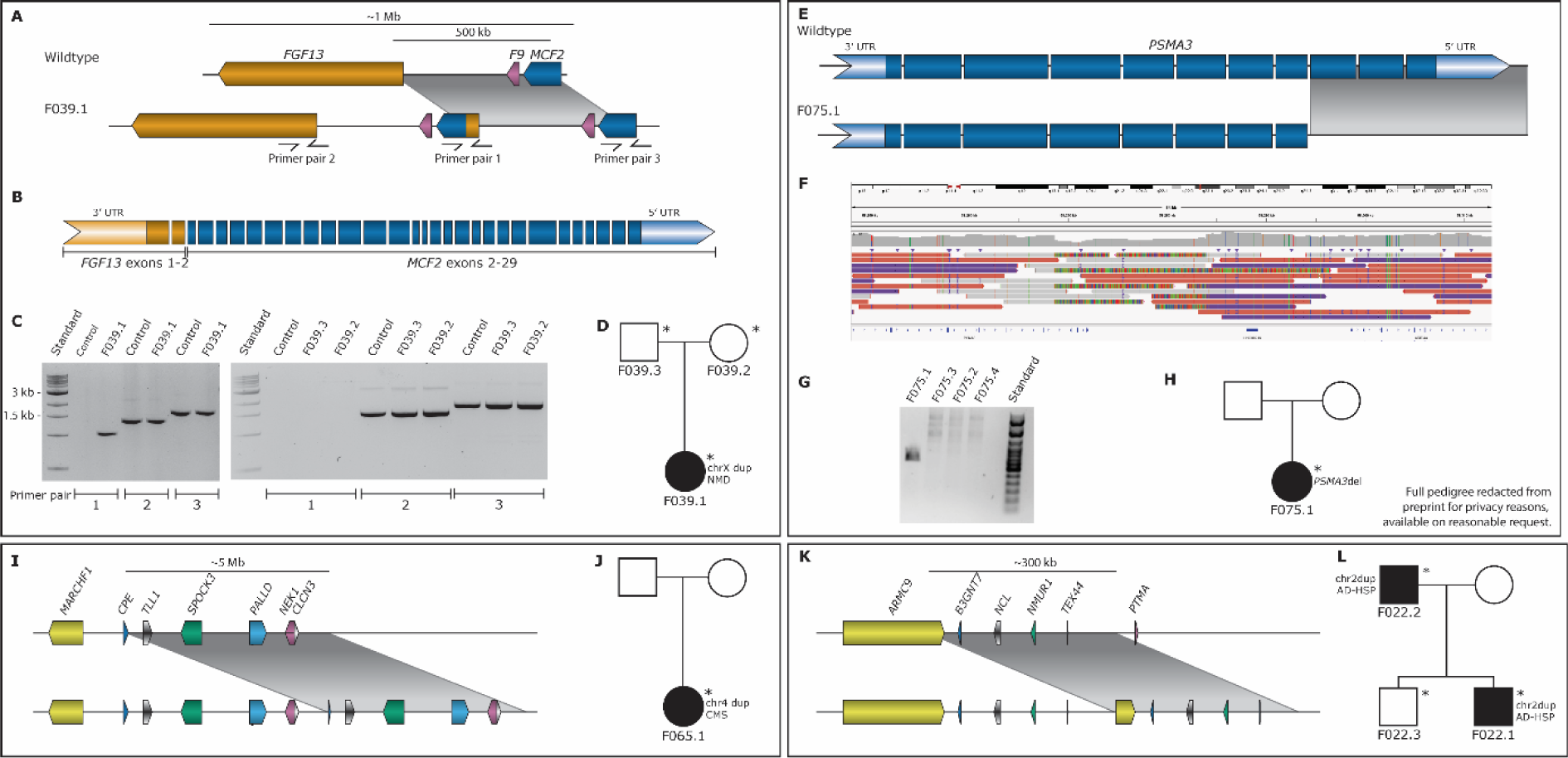
Visualization of candidate disease-causing SVs in the ‘unsolved’ cohort. In a sporadic patient (F039.1) with arthrogryposis multiplex congenita, we detected an X-chromosomal tandem duplication (A-D). The duplication spans from intron 1 of *MCF2* to intron 2 of *FGF13* and includes F9 (A). The result of the duplication is a hypothetical fusion gene including *FGF13* exons 1-2 and *MCF2* exons 2-29 (B). The duplication was validated by PCR and agarose gel electrophoresis (C) using primers targeting the breakpoints of the duplication, and a combination of the *MCF2* forward and *FGF13* reverse primers (Primer pair 1, A and C). In a sporadic patient with psychomotor development delay (F75.1) we detected a deletion of *PSMA3* exons 9 to 11, here shown as a cartoon (E) and as a screenshot using IGV (F). The deletion was validated by agarose gel electrophoresis (G) and Sanger sequencing. The index F075.1 has three healthy siblings who do not carry the deletion (H). In a sporadic female patient (F065.1) with congenital malformation syndrome, we detected a 5 Mb tandem duplication on chromosome 4, visualized here as a cartoon (I). In a sporadic male patient (F022.1) with autosomal dominant spastic paraplegia, with a similarly affected father, we detected a 300 kb tandem duplication on chromosome 2, visualized here as a cartoon (K). Sequenced individuals are marked with an asterisk (*). Abbreviations: NMD = neuromuscular disorder, CMS = congenital malformation syndrome, AD-HSP = autosomal dominant hereditary spastic paraplegia.

*FGF13* modulates the function and location of voltage-gated sodium channels in the brain (Fry et al., 2021). Mutations in the gene have been linked to developmental and epileptic encephalopathy 90 (MIM ID #301058) and intellectual developmental disorder (MIM ID #301095). *MCF2* is an oncogene belonging to a family of GDP-GRP exchange factors, the role of which is to modulate the activity of small GTPases in the Rho family. In addition to brain tissue, it has relatively high expression in the adrenal gland, the testes and the ovaries. Molinard-Chenu and colleagues reported a putative pathogenic missense mutation in *MCF2* in a patient presenting with complex perisylvian syndrome and demonstrated that murine Mcf2 controls the migration of cortical projection neurons in mice (Molinard-Chenu et al., 2020).

The duplication results in a hypothetical *FGF13*-*MCF2* fusion gene, in which the breakpoint resides within the second intron of *FGF13* and the first intron of *MCF2* (Fig. 6B). The second exon of *MCF2* is normally partly untranslated, but it is assumed that it is entirely translated in the fusion gene product; in this case, the entire fusion gene product is in-frame. The putative pathogenic mechanism of this fusion gene will be the subject of another study.

#### *PSMA3* C-terminal deletion

In a sporadic female patient (F075.1) we identified a *de novo* 15.3 kb deletion on chromosome 14 affecting the three last exons (ex 9-11) of *PSMA3* (Proteasome subunit, Alpha type, 3, MIM ID *176843) (Fig. 6E-H), chr14:58,268,649-58,283,944. The phenotype consists of psychomotor development delay, facial dysmorphism, and intellectual disability. Nerve conduction studies revealed an axonal sensorimotor neuropathy. Unaffected siblings of the index patient did not carry the deletion, and haplotyping of *PSMA3* suggests that the deletion has arisen *de novo* in the patient. The deletion breakpoints were confirmed by Sanger sequencing, and its absence in the siblings was confirmed by PCR and gel electrophoresis (Fig. 6G).

*PSMA3* is expressed in tissues throughout the body, including skeletal muscle and nerve tissues. As a proteasome subunit, the role of *PSMA3* is to contribute to the proteolytic pathway of aberrant proteins and/or proteins with high turnover rates in the ubiquitin-proteasome system (UPS). Variants in *PSMA3* have not previously been linked to disease and no structural variants only affecting *PSMA3* are described in any public databases. However, variants in genes contributing to the UPS have been linked to several neurodegenerative diseases caused by the aggregation of neurotoxic proteins in the absence of a functioning ubiquitin-proteasome system. Biran and colleagues have proposed that the *PSMA3* C-terminal region targets intrinsically disordered proteins for degradation and would thus play an important role in the ubiquitin-proteasome system (Biran et al., 2022).

#### *De novo* duplication on chromosome 4

In a singleton female patient (F065.1) with a congenital malformation syndrome, we identified a 5 Mb tandem *de novo* duplication on chromosome 4 (Fig. 6I-J). The patient presented with a complex phenotype involving growth delay, facial syndromic features with optical and neurological involvement, cleft palate, and tonic-clonic seizures. The duplicated sequence is chr4:165,447,976-170,473,341 (hg38) and involves several known disease-causing genes, amongst which *NEK1* (MIM ID *604588), and *CLCN3* (MIM ID *600580).

While none of the known disease-causing genes within the duplicated region can be directly tied back to the phenotype of the patients, some overlap is present. Variants in *NEK1* are a known cause of a form of thoracic dysplasia (short-rib thoracic dysplasia 6 with or without polydactyly, MIM ID #263520). This syndrome involves cleft palate and enlargement of the lateral ventricles; however, it is also characterized by several clinical manifestations not present in the patient. In turn, missense variants in *CLCN3* are a known cause of autosomal dominant neurodevelopmental disorder with seizures and brain abnormalities (MIM ID #619512). This disorder is characterized, among other symptoms, by dysmorphic facial features, hypertelorism, strabismus, abnormalities of the cerebellum and corpus callosum, and, in some patients, seizures.

#### 300 kb duplication on chromosome 2

In a family presenting with hereditary spastic paraplegia, we identified a 300 kb tandem duplication on chromosome 2 in the affected father (F022.2) and an affected son (F022.1) (Fig. 6K-L). The non-affected brother of the son (F022.3) does not carry the duplication. The duplicated sequence is chr2:231,348,004-231,684,006 and breaks *ARMC9* (armadillo repeat-containing protein 9, MIM ID *617612). RNA-seq confirmed the upregulation of several of the genes within the duplicated sequence.

Variants in *ARMC9* are a known cause of Joubert syndrome 30 (MIM ID #213300). Joubert syndrome is a genetically heterogeneous neurodevelopmental ciliopathy, characterized by a distinctive “molar tooth sign” brain malformation (Maria et al., 1997). Individuals with Joubert syndrome present with ataxia, along with hypotonia, abnormal eye movements, and cognitive impairment (Latour et al. 2020).

## Discussion

The Solve-RD consortium has set itself the goal to genetically diagnose previously undiagnosed rare disease families. In our current study, HiFi long-read genome sequencing was conducted for 293 carefully selected patients and healthy relatives from 114 rare disease families. The cohort was further divided into two subcohorts, consisting of ‘unsolvable’ syndromes in which we expect the molecular cause to be currently unknown, and ‘unsolved’ families with RNDs or NMDs in which we expect to identify new variants in known or novel disease genes.

Whereas sequencing was performed at a relatively modest coverage of ∼10-fold, we identified and orthogonally validated pathogenic variants of all classes: SNVs, InDels, SVs, and STRs. Although our approach is not ideally suited for obtaining highly accurate SNV call sets, strict filtering, interpretation, and validation of calls show that previously unidentified and/or misclassified SNVs also contribute to the diagnostic yield of our study. For example, we identified a previously unidentified deep-intronic hemizygous *DMD* variant leading to altered splicing in a male patient with muscular dystrophy, as well as an intronic *SPAST* variant segregating in a family with hereditary spastic paraplegia, which was not considered for clinical interpretation in the initial exome analysis.

In two cases, our study did not identify a novel variant but provided additional information about previously identified candidate variants. In the case of a titinopathy patient, a pathogenic maternally inherited mutation, and a single base pair *de novo* deletion in *TTN* had already been identified. However, the diagnosis was inconclusive because the allele on which the *de novo* mutation had occurred could not be determined. The long reads of this study allowed for the phasing of this variant and could confirm that it occurred on the paternal allele thereby leading to a definite diagnosis. Similarly, a previously detected gain involving *MYOT* was shown here to be a tandem duplication.

In total, we identified disease-causing variants in 12 out of 93 families with unsolved disorders (13.0%), but only one in the 21 families (4.8%) with ‘unsolvable’ disorders. The variant found in the ‘unsolvable’ trio is a *de novo* SNV in *TUBA1A* in a patient initially suspected of Aicardi syndrome. The identified *TUBA1A* variant has previously been associated with LIS3 (Bahi-Buisson et al., 2014b). In hindsight, and by reverse phenotyping, the clinical experts in this project also confirmed this as the disease-causing genetic variant in this specific case. Individuals with LIS have severe neurological problems, including intellectual disability and epilepsy, and may appear phenotypically very similar to patients with Aicardi syndrome. The difference in the number of resolved cases between the two cohorts suggests that ‘unsolvable’ syndromes are indeed a special class of syndromes. In such cohorts, other explanations for the disorder should be considered, such as methylation defects, somatic mutations, polygenic origin, larger heterogeneity than expected, or even non-genetic causes of disease (Boycott et al., 2018).

Next to the 13 diagnoses we also identified four candidate disease-causing SVs: one intragenic deletion in *PSMA3*, two large duplications (a 5Mb event breaking and involving multiple coding genes, and a 300kb event affecting the *ARMC9* gene) and an X-chromosomal duplication likely leading to the production of a *FGF13*-*MCF2* fusion protein.

Of these, two events, the intragenic deletion in *PSMA3* and the X-chromosomal duplication were *de novo* events. Although for *PSMA3,* parental samples were not available, we were able to infer the *de novo* status of the deletion by using the long reads to reconstruct the haplotype on which the deletion occurred and observing that the deletion was not present in other siblings with the same haplotype. The low SV mutation rate in humans (strong evolutionary constraint against SVs) implies that such genic *de novo* SV events are good candidates for pathogenicity (Porubsky & Eichler, 2024).

The *de novo* deletion affects the last three exons of *PSMA3* in a sporadic patient with a phenotype similar to Charcot-Marie-Tooth disease type 2. The deletion likely results in truncated mRNA and subsequent nonsense-mediated decay. Mutations in *PSMA3* have not been described in literature, however, the gene encodes for a subunit of the ubiquitin-proteasome system (UPS). Variants in other genes encoding for UPS subunits have been described as causative in several neurodegenerative disorders; in these, the absence of a functioning ubiquitin-proteasome system leads to the aggregation of neurotoxic proteins. Moreover, the gene is highly intolerant to loss-of-function variants, making it a likely candidate for a dominant disease gene. *PSMA3* is (lowly) expressed in whole blood and future gene expression analysis in patient material would likely provide further support for the disease-causing nature of the deletion.

The *de novo* duplication on the X-chromosome of a patient with arthrogryposis multiplex congenita, thoracolumbar scoliosis, and restrictive ventilatory defect likely results in a fusion *FGF13-MCF2* gene. *FGF13* is an X-linked dominant disease gene associated with neurodevelopmental disorder phenotypes, different from the phenotype observed in this female patient. *F9* is not known to be associated with a disease once duplicated, and also *MCF2* has not been associated with disease yet. We can hypothesize that the creation of an *FGF13-MCF2* fusion gene and especially the simultaneous loss of an *FGF13* wildtype allele may have a phenotypic consequence for this patient. However, further investigations, including X chromosome inactivation and functional studies, will be needed to understand the relationship between this event and the patient’s phenotype.

Out of the four candidate variants, three concern variants affecting already established disease genes. Confirmation of the pathogenic nature of these variants may broaden the known phenotypic spectrum of the affected genes or establish new inheritance patterns. Only the *de novo* deletion affecting *PSMA3* suggests a potential novel neurodevelopmental disease gene.

In our study, the clinical interpretation of variants was hampered by the large number of identified “rare” SV. Large catalogues of identified variants from long-read sequencing of both affected and unaffected individuals will therefore be of critical importance to improve variant interpretation in such cases. Control population efforts are underway, for example in the All of Us Research Program (Mahmoud et al., 2024) or initiatives such as colors-db (https://colorsdb.org/). Such control cohorts may, in the future, help to diagnose additional patients in our cohort. Here, Solve-RD shares the full dataset, including expert-curated pedigree and phenotype information (EGA: accession number pending). In addition, we also share a frequency call set of high-quality SVs of the unrelated individuals as a resource for other researchers (Methods; EGA: accession number pending). This resource shall prove valuable in particular with the increase in novel variant types for which LRS has higher sensitivity.

Now that whole genome LRS is becoming more affordable for large-scale studies like ours, and can be performed at higher coverages, we expect many new (hidden) variant discoveries in the future. The fact that some relevant variants (in for example the *MYOT* case) were only identified through visual inspection of the sequencing data, suggests that improvements in variant calling algorithms may also contribute significantly to the resolution of additional cases (Ebert et al., 2021).

In conclusion, HiFi long-read genome sequencing was conducted for a unique cohort of 293 individuals from 114 previously studied rare disease families. While we did not identify a common genetic cause in any of the ‘unsolvable’ syndromes, we identified causal genetic variants in 13.0% of families from the ‘unsolved’ cohort, and candidate variants in an additional 4.3%. Our study shows the potential and effectiveness of even modest coverage LRS in rare disease studies.

## Methods

### Study cohort

HiFi long-read genome sequencing was conducted for 293 individuals from 114 genetically undiagnosed rare disease families (Supplemental Table S1). Patient samples came from two sub-cohorts: the ‘unsolvables’ (n=61) for which genetic causes remain unknown, and the ‘unsolved’ (n=232) for which a previously hidden genetic variant in a known or yet unknown disease gene is expected to be the cause of disease. All of the patients and healthy relatives were carefully selected by experts from four European Reference Networks: RND (38 families; 95 individuals), EURO-NMD (37 families; 89 individuals), ITHACA (32 families; 88 individuals; including all ‘unsolvables’), and EpiCARE (7 families; 21 individuals). Depending on the research hypothesis and sample availability 1 to 7 (un)affected individuals were selected per family for sequencing on a PacBio Sequel IIe instrument. The most represented family structure is the parent-offspring trio (n_families_= 42; n_samples_ = 126; 43.0% of cohort). We have used a single SMRT cell of sequencing data per individual which, after read alignment (onto GRCh38) and read filtering, resulted in a mean HiFi read depth of 9.8 (Supplemental Table S4).

### DNA sequencing

Genomic DNA was isolated from peripheral blood according to standard protocol and long-read genome HiFi sequenced using SMRT sequencing technology (Pacific Biosciences, Menlo Park, CA, USA). For every sample, 7-15 µg of DNA was sheared on Megaruptor 2 or 3 (Diagenode, Liège, Belgium) to a target size of 15-18 kb. Libraries were prepared with SMRTbell Prep Kit 2.0 or 3.0. Size selection was performed using a BluePippin DNA size selection system (Sage Science, Beverly, MA, USA) targeting fragments equal to or longer than 10 kb in length. Sequence primer and polymerase were bound to the complex using the Sequel II binding kit 3.2 (PacBio), and sequencing was performed on the Sequel IIe system with 2.0 Chemistry and 30h movie time per SMRTcell using a single flow cell per sample.

### Primary data analysis

All samples were processed in the same fashion using a custom workflow based on standard methods from the Pacific Biosciences analysis pipeline (https://github.com/PacificBiosciences/pb-human-wgs-workflow-snakemake) (Supplemental Fig. S2). Sequencing reads were aligned to the GRCh38/Hg38 genome with pbmm2 (version 1.4.0) (Li, 2018, 2021), using default parameters. HiFi reads (>QV20) were extracted for all downstream analyzes. Small variant (substitution and indel) calling was performed using DeepVariant (version 1.1.0) with default settings (Poplin et al., 2018). No threshold for the maximum size of the indels was applied, and all indel calls were used for further analyses. For parent-offspring trios, GLNexus (version 1.3.1) was used to conduct SNV joint genotyping (Yun et al., 2021).

Small variants were phased using Whatshap (version 1.1.0) and variants were annotated using an in-house developed pipeline (Martin et al., 2016). This variant annotation was based on the Variant Effect Predictor (VEP V.91) and Gencode 34 basic gene annotations. STR calling was performed using Tandem Repeat Genotyper (TRGT; version 0.3.3) at 56 known disease-associated STR loci (Dolzhenko et al., 2024; Supplemental Table S5). SV calling was performed using PBSV (version 2.4.0) using default settings with a minimum SV size of 20 bp (https://github.com/PacificBiosciences/pbbioconda). SVs were annotated using AnnotSV (version 3.1.1; (Geoffroy et al., 2018).

In each of the 114 RD families that comprise our study cohort, we selected the maximum number of unrelated individuals resulting in a subcohort of 166 unrelated individuals. SVs merging using Jasmine resulted in a call set of 251,672 unique SVs (corresponding to 11,290,783 variant alleles in the subcohort) of which 59,876 are private to one individual (Kirsche et al., 2023). Only 1,971 unique SVs (0.78%; 51,433 alleles) in the complete call set affect a coding exon. An additional 2,965 unique SVs (1.18%; 111,231 alleles) alter the non-coding sequence of an exon and 95,197 unique SVs (37.8%; 4,445,817 alleles) reside in an intron of a protein-coding gene. Lastly, 35,525 unique SVs (14.1%; 1,638,574 alleles) affect a non-coding gene.

The distribution of sequence gains and losses in our study cohort is characterized, as expected, by the inverse relation between SV length and frequency. Exceptions to this smooth decrease in density are the characteristic peaks for short interspersed nuclear elements (SINE) and long interspersed nuclear elements (LINE) peaks which are respectively present at medians +/− 323 bp and +/− 6,050 bp (Supplemental Fig. S3).

### Variant filtering

#### Structural Variants

In parent-offspring trios, we focused on putative *de novo* variants. For this, we selected sites which are covered by at least 8 HiFi reads in each of the members of the trio. Furthermore, at least 3 HiFi reads should support the variant allele in the child. Because of the modest sequencing depth, we subjected all of the resulting SVs to visual inspection using IGV. In this step, we removed SV calls that were unclear in the child (despite the variant call), SVs for which one of the parents had a trace in their sequencing data (for example supported by one read) and SVs for which both or one of the parents only had 1 allele sequenced (based on the phased alignments). All the remaining sites were subjected to primer design for further validation (cf. wet-lab validation).

In all other family structures, we focused on rare inherited high-quality SVs that co-segregate with disease. To do so, we selected family-unique SV calls which were observed in all affected members of a given family and absent from all unaffected family members. Furthermore, in at least one of the affected family members the SVs need to be covered by at least 8 HiFi reads of which 3 support the variant allele. In contrast to SV calls corresponding to well-characterized deletions, inversions, duplications, and insertions we visually inspected all break end-calls. We evaluated, based on coverage and the complexity of the sequence context, whether a break end-call could, together with the linked break end-call, be a signature of a genetic event that is too large to be characterized as a deletion, inversion, duplication or insertion by pbsv. In this step, we required that all clipped reads support the same regional split. Since these calls support relatively large genetic events (> 100 kb) we clinically assessed them in the complete human genome. In contrast, clinical interpretation of SV calls corresponding to characterized deletions, inversions, duplications, and insertions (size < 100 kb) was restricted to events that reside in genes within ERN-specific gene lists (S. Laurie, W. Steyaert, E. de Boer et al., Nat Med in revision).

#### Single nucleotide variants

In parent-offspring trios, we focused on putative de novo events. These were selected from the joint calls generated by GLnexus. We considered a variant to be putative *de novo* when the child is heterozygous (genotype ‘0/1’) and both parents are homozygous for the reference allele (genotype ‘0/0’). In addition, we require that both parents and the child have ≥ 8 HiFi reads covering the site of which 3 reads support the variant allele in the child.

In all other families, we selected for rare inherited SNVs that co-segregate with disease. For this, we selected SNVs that are unique to a single family that are present in all affected family members and absent from all unaffected family members. In contrast to the *de novo* variant interpretation, we restricted variant interpretation for inherited variants to variants that reside in genes incorporated in the ERN-specific gene lists.

#### STR genotypes

STR genotypes were visualized in R per submitter group in comparison with the rest of the cohort to facilitate the evaluation of the quality of calling per loci and the detection of pathogenically expanded alleles. These results were sent to the groups for clinical interpretation.

### Wet-lab variant validation

Primers for the validations were designed using the online Primer3 design tool as per the manufacturer’s suggestions. Primers were selected to be 18-21 nucleotides in length with a GC-content ranging from 40-60%. While an annealing temperature of 60°C was proposed to be optimal, annealing temperatures between 57-61°C were considered acceptable as well. Sizes of the products ranged between 1000-4000 nucleotides, to ensure capture of the full region and compatibility with PacBio LRS.

In three cases, two adjacent SVs could be covered by one primer pair, and for the large X-chromosomal duplication, altogether three primer pairs were designed (Fig. 6). For 8 variants, primer design was not possible. In addition, primers were designed for the candidate exon 6 deletion in *REEP1* and a 50 bp deletion in *MAPK8IP3* segregating with disease in F008.1 and F008.2 (Supplemental Table S3).

The running conditions for the remaining 26 primer pairs were optimized in gradient runs using melting temperatures of 60, 61, and 62°C and extension times appropriate to product length using LongAmp HotStart Taq 2x MasterMix (New England Biolabs, Ipswich, MA, USA). The primer pairs were optimized successfully for 18 candidate *de novo* variants.

All optimized primer pairs were run on patient, control, and parental DNA. The samples were cycled as follows: 94°C 30 seconds; 27 cycles of 94°C 30 seconds, 60-62°C 1 minute, 65°C 1 minute 40 seconds (short), 3 minutes 20 seconds (long) or 5 minutes (ultra); 65°C 10 minutes; 4°C hold. The short program was used for amplicons under 2500 bp, the long program for amplicons between 2500 and 3500 bp, and the ultra-long program for amplicons over 6000 bp. The PCR products were verified by agarose gel electrophoresis.

All successfully amplified patient samples were validated by targeted LRS. Subsequent sequencing of parental samples was performed as per the workflow above for samples in which the variant call was confirmed in the index.

## Data access

Long-read sequencing data for the complete study cohort have been deposited in EGA under control access (accession codes).

## Data Availability

Long-read sequencing data for the complete study cohort have been deposited in EGA under controlled access (accession codes pending).

## Acknowledgements

The Solve-RD consortium is grateful to all involved RD patients and their families as well as other contributors to Solve-RD.

The Solve-RD project has received funding from the European Union’s Horizon 2020 research and innovation program under grant agreement No 779257. This research is supported (not financially) by several ERNs: ERN on Intellectual disability, TeleHealth, Autism and Congenital Anomalies (ERN ITHACA)—Project ID No 101085231; ERN on Rare Neurological Diseases (ERN RND)—Project ID No 101155994; ERN for Neuromuscular Diseases (ERN Euro-NMD)—Project ID No 101156434; and ERN for Rare and Complex Epilepsies (ERN EpiCARE) - Project ID No 101156811. The ERNs are co-funded by the European Union within the framework of the Third Health Program. We would also like to thank all other Solve-RD colleagues who were not mentioned by name in the author list, including members of the Solve-RD data interpretation task force (DATF), and other members of ERNs and DITFs.

We also thank The Radboud Technology Center Genomics for the library preparation and sequencing of all samples.

V.A.Y. and J.G. received funding from the Deutsche Forschungsgemeinschaft (DFG, German Research Foundation) via the project NFDI 1/1 “GHGA - German Human Genome-Phenome Archive” (No 441914366).

L.Sa. received funding from the Sigrid Jusélius Foundation (Fellowship No 220540).

R.S. received funding from the Bundesministerium für Bildung und Forschung (BMBF) through funding for the TreatHSP network (grant 01GM2209A) and the National Institute of Neurological Disorders and Stroke (NINDS) under Award Number R01NS072248.

H.H. was supported by the DFG (HE8803/1–1 to H.H.)

