## Supplemental Figures S1-S3, Supplemental Table S1-S5 captions for "Unravelling undiagnosed rare disease cases by HiFi long-read genome sequencing"

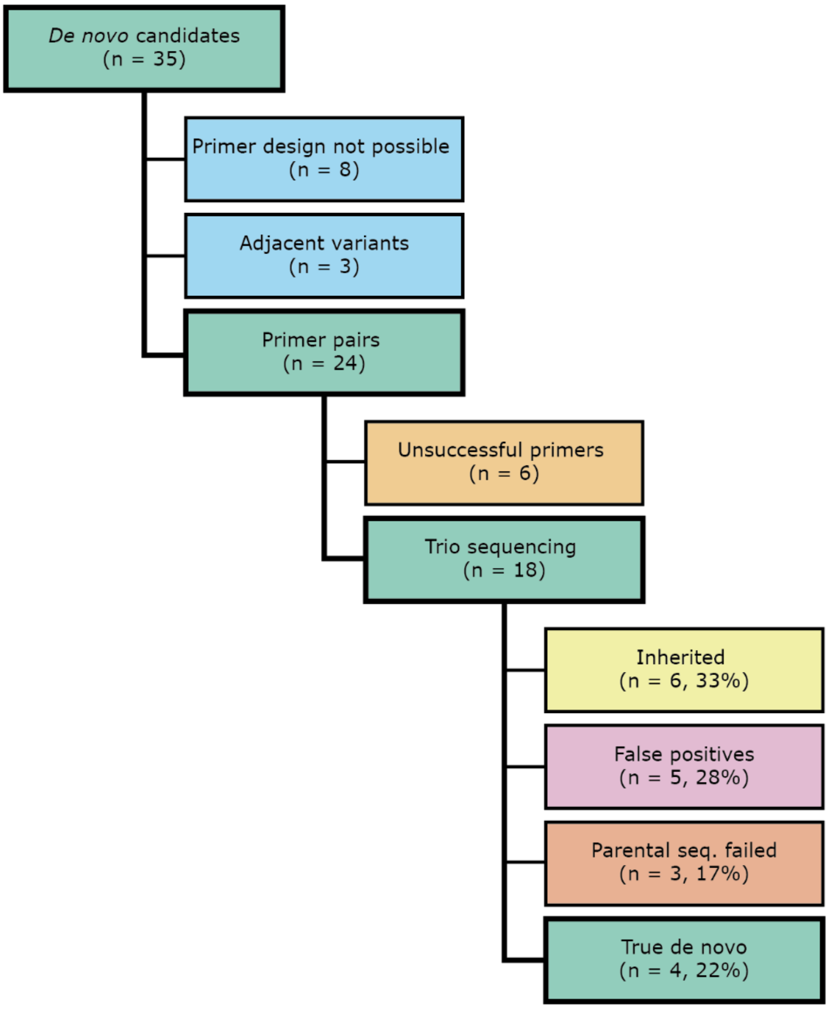


**Figure S1**: Flow of de novo variant validations by targeted LRS.

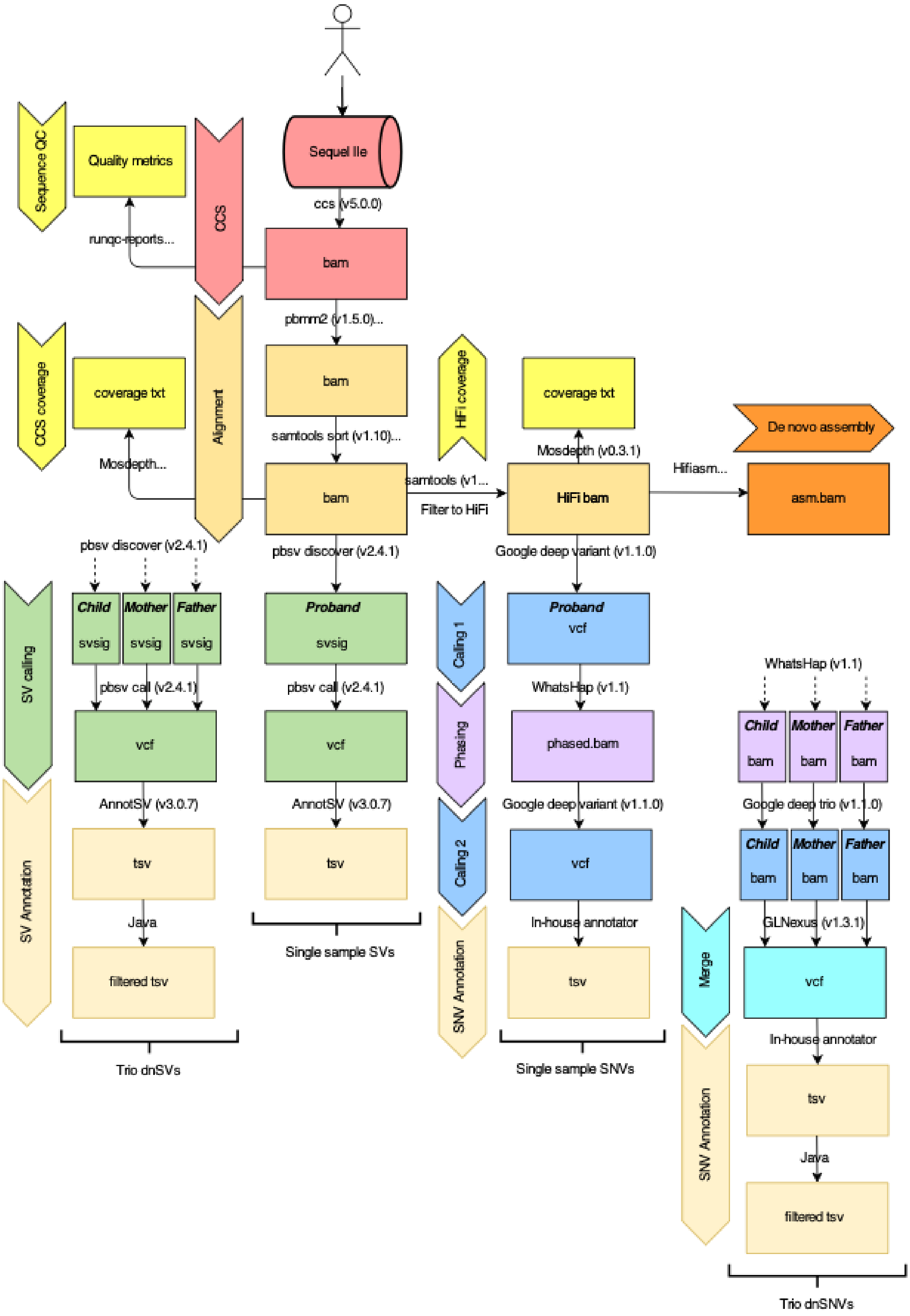


**Figure S2:** Schematic overview of the analysis workflow

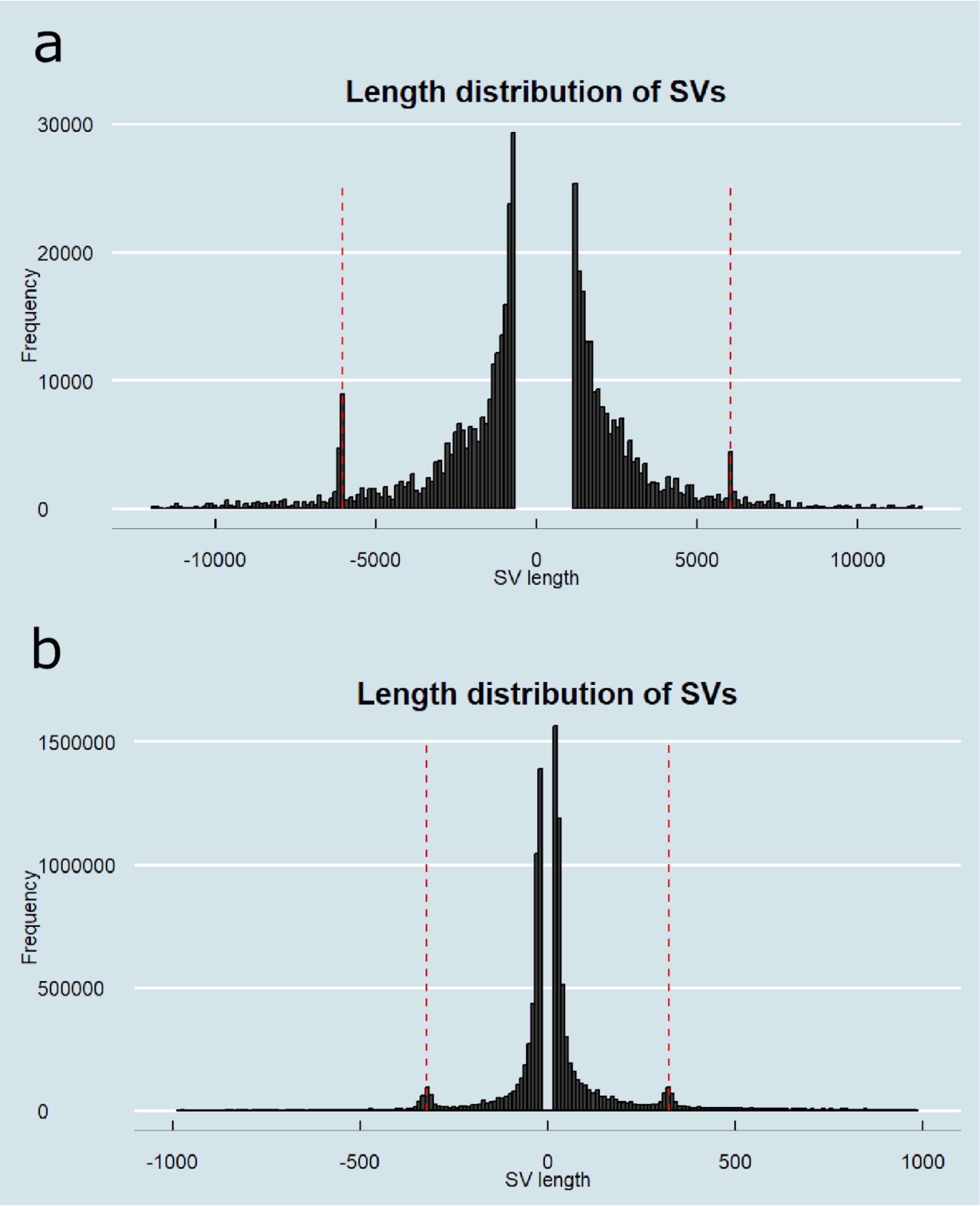


**Figure S3:** SV length distribution for sequence gains (insertions and tandem duplications) and sequence losses (deletions) visualized using different length and frequency ranges. In panel a the full length spectrum of sequence losses and sequence gains is visualized. In this plot, the peaks corresponding to the long interspersed nuclear elements become apparent at plus and minus 6,050 bp. Similarly in panel b where the horizontal axis ranges from -1,000 to +1,000 bp, the peaks corresponding to the short interspersed nuclear elements become apparent at 323 bp.

### Supplementary Table Captions

Table S1: Overview of study cohort. The participant ID can be found in column A. Column B indicates the subcohort (‘unsolvables’ or ‘unsolved’) to which the participant belongs. Gender, family identifier, Ern and Disease status ((A)ffected or (N)ot (A)ffected) can respectively be found in columns C, D, E and F.

Table S2: Number of variant calls for all participants in the study cohort. Abbreviations: BND: break end calls, DEL: deletions, INV: inversions, DUP: duplications, INS: insertions. Last 2 rows in the table summarize the data by their median and mean.

Table S3: Overview of validations for which PCR primers were designed. 21 pairs of primers were successfully designed for validation of putative *de novo* variants. In addition to these variants we validated 4 inherited variants. Column H indicates the genomic location of the variant relative to hg38.

Table S4: Overview of the HiFi sequencing coverage for the complete study cohort. Fraction_X refers to the fraction of the genome that is covered by at least X reads. Column N indicates the mean genome-wide HiFi read depth.

Table S5: Overview of the 56 STR loci that were genotypes in this study.
